# Sexual and Relationship Content Exposure on Social Media and Adolescent Sexual Health Attitudes

**DOI:** 10.64898/2026.11.12.26367956

**Authors:** Zhiying Yue, Michael Rich, David Bickham

## Abstract

**Purpose:** To identify patterns of adolescents’ reported exposure to sexual and relationship content on social media and examine their associations with permissive sexual attitudes and perceived barriers to condom use.

**Methods:** In a cross-sectional online survey (January to February 2026), 1,124 US high school students aged 16 to 18 years rated their typical exposure to 20 content categories developed with Youth Advisory Board input. Network analysis identified content communities, latent profile analysis grouped adolescents by community-level exposure, and covariate-adjusted linear models compared attitudes across profiles.

**Results:** Four content communities and 4 exposure profiles were identified. Adolescents in the High Overall Exposure profile (n = 299; 27.3%) reported frequent exposure across all content communities and had higher adjusted permissive sexual attitudes (difference, 0.58; 95% CI, 0.40 to 0.75; d = 0.69) and perceived condom-use barriers (difference, 0.50; 95% CI, 0.31 to 0.69; d = 0.61) than the Moderate General Exposure profile. Differences were attenuated but persisted after adjustment for sociodemographic characteristics, sensation seeking, religiosity, and social media time (0.39 and 0.38) and were robust to data-quality exclusions and correction for classification error.

**Discussion:** Adolescents’ exposure to sexual and relationship content on social media is patterned, and higher overall exposure is associated with attitudes relevant to condom use. Sexual health counseling may benefit from addressing the broader mix of messages adolescents encounter online.

**Implications and Contribution:** Adolescents’ reported exposure to sexual and relationship content on social media formed distinct patterns. Higher overall exposure, particularly to sexual rather than relationship-focused content, was associated with more permissive sexual attitudes and greater perceived condom-use barriers, suggesting that sexual health counseling should consider adolescents’ online content environments.

## Introduction

Adolescents’ social media use is intensive and increasingly personalized [1]. In a 2025 Pew Research Center survey of US teens aged 13 to 17 years, approximately 1 in 5 reported using TikTok or YouTube almost constantly [2]. A 2023 Gallup survey estimated that adolescents spent an average of 4.8 hours per day across seven social media platforms [3]. National concern about youth social media use has prominently focused on mental health [4]. Yet social media also provide an everyday context for adolescent sexual development, where young people encounter messages about dating, intimacy, attractiveness, sexual behavior, and condom use [5].

These media experiences unfold against persistent challenges in adolescent sexual health. In 2023, people aged 15 to 24 years accounted for 48% of reported US cases of chlamydia, gonorrhea, and syphilis [6]. Correct and consistent condom use reduces the risk of HIV and other sexually transmitted infections [7], yet only 52% of currently sexually active high school students reported using a condom at last sexual intercourse in 2023 [8]. A meta-analysis of 249 studies found that adolescents’ attitudes, intentions, and partner communication were associated with condom use [9]. These findings motivate examining sexual health attitudes and perceived condom-use barriers in relation to sexual and relationship messages encountered through social media.

Social media may contribute to adolescent sexual socialization through the messages adolescents encounter and the visible audience responses those messages receive. Posts about dating, relationship expectations, sexualized self-presentation, and protection may communicate messages about what is typical, desirable, or socially acceptable [10,11]. Sexual script theory emphasizes how such messages provide frameworks for interpreting sexual situations, while social cognitive theory highlights learning through observation of models, their consequences, and others’ responses [12,13]. Consistent with these frameworks, a meta-analysis linked sexual media exposure with sexual attitudes, perceived peer norms, and sexual behavior, with stronger associations among adolescents than emerging adults [14].

The personalized and interactive features of social media add another dimension to these socialization processes. Recommendation systems shape subsequent content based on viewing and engagement, and likes, comments, and view counts signal audience responses [15].

Adolescents may therefore differ in which kinds of sexual and relationship content they see. Existing research has estimated cumulative sexual media exposure [16] or examined sexual messages in youth-oriented media [17,18], but has not characterized how adolescents’ exposure to different categories of sexual and relationship content on social media is patterned.

Using a 20-item exposure measure developed with input from adolescent members of a Youth Advisory Board, we examined sexual and relationship content exposure among US adolescents aged 16 to 18 years. Network analysis characterized conditional associations among content-category exposure ratings, while latent profile analysis identified groups of adolescents with different configurations of community-level exposure scores. We then examined associations between these exposure profiles and sexual health attitudes and perceived barriers to condom use, and tested whether associations reflected how often adolescents encountered such content overall or the particular mix of content they reported.

## Methods

### Study Design and Participants

Data were obtained from a cross-sectional online survey of US adolescents aged 16 to 18 years conducted from January through February 2026. Participants were recruited through Alchemer, which routed potential participants from national online survey panels. Eligible participants were high school students residing in the United States. Quota targets were established before data collection to approximate US distributions of age, gender identity, race and ethnicity, and school type; data were not weighted. Of 2,779 individuals routed to the survey, 2,699 entered screening, 1,854 met eligibility criteria, and 1,125 completed the survey. Participant characteristics are shown in Table 1.

**Table 1.** Participant Characteristics, Overall and by Exposure Profile.

| **Characteristic** | **Total (N = 1,124)** | **Relationship-Socialization (n = 284)** | **Moderate General (n = 314)** | **High Overall (n = 299)** | **Low-Intensity Relationship (n = 199)** | **P value** |
|---|---|---|---|---|---|---|
| Age, mean (SD), y | 17.11 (0.79) | 17.07 (0.81) | 17.21 (0.80) | 17.10 (0.77) | 16.99 (0.76) | .01 |
| Gender identity |  |  |  |  |  | <.001 |
| Female | 565 (50.3) | 169 (59.5) | 168 (53.5) | 137 (45.8) | 79 (39.7) |  |
| Male | 515 (45.8) | 102 (35.9) | 135 (43.0) | 153 (51.2) | 112 (56.3) |  |
| Nonbinary or another identity | 35 (3.1) | 10 (3.5) | 10 (3.2) | 7 (2.3) | 7 (3.5) |  |
| Sexual minority | 281 (25.0) | 71 (25.0) | 83 (26.4) | 68 (22.7) | 54 (27.1) | .61 |
| Transgender identity | 58 (5.2) | 13 (4.6) | 19 (6.1) | 10 (3.3) | 16 (8.0) |  |
| Race and ethnicity |  |  |  |  |  | .02 |
| Hispanic or Latino (any race) | 245 (21.8) | 63 (22.2) | 66 (21.0) | 71 (23.7) | 36 (18.1) |  |
| White | 575 (51.2) | 152 (53.5) | 146 (46.5) | 153 (51.2) | 112 (56.3) |  |
| Black | 141 (12.5) | 24 (8.5) | 55 (17.5) | 40 (13.4) | 19 (9.5) |  |
| Asian | 60 (5.3) | 24 (8.5) | 16 (5.1) | 8 (2.7) | 11 (5.5) |  |
| Another race or multiple races | 97 (8.6) | 19 (6.7) | 30 (9.6) | 26 (8.7) | 21 (10.6) |  |
| Not reported | 6 (0.5) | 2 (0.7) | 1 (0.3) | 1 (0.3) | 0 (0.0) |  |
| School type |  |  |  |  |  | <.001 |
| Traditional public | 782 (69.6) | 219 (77.1) | 240 (76.4) | 155 (51.8) | 146 (73.4) |  |
| Charter, magnet, or vocational | 182 (16.2) | 35 (12.3) | 32 (10.2) | 94 (31.4) | 19 (9.5) |  |
| Private, homeschool, or other | 160 (14.2) | 30 (10.6) | 42 (13.4) | 50 (16.7) | 34 (17.1) |  |
| Ever had sex |  |  |  |  |  | <.001 |
| Yes | 349 (31.0) | 66 (23.2) | 112 (35.7) | 132 (44.1) | 31 (15.6) |  |
| No | 683 (60.8) | 192 (67.6) | 178 (56.7) | 153 (51.2) | 151 (75.9) |  |
| Prefer not to answer or missing | 92 (8.2) | 26 (9.2) | 24 (7.6) | 14 (4.7) | 17 (8.5) |  |
| Currently dating or talking to someone | 541 (48.1) | 124 (43.7) | 155 (49.4) | 182 (60.9) | 68 (34.2) | <.001 |
| Daily hours on YouTube, Instagram, and TikTok, mean (SD) | 6.6 (3.6) | 6.4 (3.8) | 6.9 (3.7) | 7.6 (3.0) | 4.8 (3.4) | <.001 |
| Sensation seeking (1-5), mean (SD) | 3.15 (0.76) | 2.93 (0.79) | 3.13 (0.67) | 3.54 (0.68) | 2.90 (0.77) | <.001 |
| Religiosity (0-5), mean (SD) | 2.32 (1.88) | 2.62 (1.83) | 2.44 (1.81) | 1.86 (1.84) | 2.33 (1.99) | <.001 |
| Not sure or not applicable to all condom-barrier items | 278 (24.7) | 83 (29.2) | 61 (19.4) | 25 (8.4) | 98 (49.2) | <.001 |

Data are No. (%) unless otherwise indicated. The total column includes all 1,124 participants; profile columns include the 1096 adolescents with valid scores for all 4 content communities. Percentages use column totals as denominators. Race and ethnicity was assessed using a select-all-that-apply format and categorized as Hispanic or Latino (any race), non-Hispanic single-race groups, and another race or multiple races. Sexual minority includes gay or lesbian, bisexual, pansexual, queer, asexual, another sexual identity, and questioning responses. Religiosity ranged from 0 (no religious affiliation) to 5 (very close identification with one's religion); sensation seeking was assessed with the Brief Sensation Seeking Scale. P values compare profiles using χ<sup>2</sup> tests or 1-way analysis of variance, excluding missing responses.

The Institutional Review Board of Boston Children’s Hospital approved this study under an anonymous, minimal-risk protocol. Parental permission was waived under the approved protocol. Participants received compensation through their panel providers. Two attention-check items were embedded in the survey, and responses failing either check were excluded before the final sample was assembled. One completed response was excluded after Alchemer’s automated data-quality system flagged it for potential speeding, straightlining, or duplication, yielding a final sample of 1,124 participants.

### Measures

#### Sexual and relationship content exposure

Exposure was assessed with a 20-item measure developed in three stages. First, the research team reviewed existing measures of sexual media exposure and sexual learning to develop an initial pool of items [19,20]. Second, trained staff conducted structured searches on TikTok and Instagram using contemporary terms related to sex, dating, and relationships to identify additional content themes. Third, Youth Advisory Board members reviewed candidate items for comprehensibility, cultural relevance, developmental appropriateness, and familiarity. Participants rated how frequently they typically encountered each category, from 1 (never) to 5 (almost always); “prefer not to answer/I don’t remember” was treated as missing (Cronbach α = .94). After each of 5 item blocks, participants reported whether such content came mostly from accounts they followed or from recommended accounts (eTable 1 in the Supplement).

Permissive sexual attitudes were assessed with a 4-item composite of endorsement of one-night stands, multiple sexual partners, sexual exchange, and condomless sex (Cronbach α = .76). Perceived barriers to condom use were assessed with 4 items (Cronbach α = .79) adapted from the Condom Barriers and Motivations Scale [21], assessing beliefs that condoms reduce sensation, disrupt the moment, feel uncomfortable, or make sex feel less intimate. Items were rated from 1 (strongly disagree) to 5 (strongly agree); barrier items also offered a “not sure/not applicable” option, treated as missing. Valid item responses were averaged, with at least 1 valid response required.

Participants reported age, gender identity, sexual orientation, transgender identity, race and ethnicity (select all that apply), school type, and whether they had ever had oral, vaginal, or anal sex with another person. Sexual minority status included gay or lesbian, bisexual, pansexual, queer, asexual, questioning, or another identity. Additional covariates were parental education, perceived family financial status, religiosity (0, no religious affiliation, to 5, very close identification with one’s religion), sensation seeking (Brief Sensation Seeking Scale; α = .79) [22], typical daily time on YouTube, Instagram, and TikTok, and current dating status.

### Statistical Analysis

Analyses were conducted in R version 4.5.1 and Python version 3.13. A regularized partial correlation network of the 20 exposure items was estimated using the extended Bayesian information criterion graphical least absolute shrinkage and selection operator (EBICglasso; γ = 0.5) with Pearson correlations [23]. Edges represented conditional associations between exposure ratings after accounting for the remaining items. Walktrap community detection [24] identified more strongly interconnected categories. Strength centrality was defined as the sum of absolute edge weights for each category; its stability was evaluated with 1,000 case-dropping bootstrap iterations [25]. Community stability was evaluated in 500 nonparametric bootstrap samples [26], and a polychoric-correlation network was estimated as a sensitivity analysis.

Community-level exposure composites were created by averaging items within each network-derived community, standardized, and entered as indicators in latent profile analysis using mclust [27]. Models with 1 to 7 profiles were estimated with equal within-profile variances and zero covariances, using 100 random starts and a strict convergence criterion. Solution selection considered the Bayesian information criterion (BIC), the bootstrap likelihood ratio test [28], entropy, average posterior probabilities, minimum profile size, and interpretability [29]. Profiles were labeled by examining mean exposure frequencies (Figure 2A) and the relative emphasis of each content community within profiles (Figure 2B).

Adolescents were assigned to their most likely profile, and linear models (equivalent to analysis of covariance) compared permissive sexual attitudes and perceived barriers to condom use across profiles, adjusting for age, a binary gender covariate, and sexual minority status. Estimated marginal means, Tukey-adjusted pairwise contrasts with simultaneous 95% CIs, and standardized differences (Cohen d; contrast divided by the residual SD) were calculated.

Sensitivity analyses sequentially added sociodemographic characteristics (race and ethnicity, school type, parental education, family finances); religiosity, sensation seeking, and social media time; and sexual experience and dating status. To account for uncertainty in profile assignment, models were re-estimated with bias-adjusted three-step (BCH) weights [30,31] and among adolescents with posterior probabilities of .80 or higher. Possible careless responding was flagged using response-pattern and consistency indicators [32] (eMethods in the Supplement); 44 adolescents (3.9%) were flagged and excluded in sensitivity analyses. Other analyses coded gender identity categorically (retaining nonbinary adolescents), required complete outcome scales, and restricted analyses to sexually experienced adolescents.

To test whether associations reflected overall exposure frequency or the content mix, each outcome was regressed on overall exposure (mean of the 4 standardized community scores) and, separately, on the 4 community scores simultaneously; the models were compared with an F test. Logistic regression examined whether profiles differed in answering “not sure/not applicable” to all 4 condom-barrier items.

## Results

### Network Structure of Content Exposure

The regularized partial correlation network of the 20 exposure-item ratings (n = 965) identified 4 content communities (modularity = 0.28): Sexual Scripting and Health Narratives, Romantic Relationship Socialization, Sexualized Appearance and Performance, and Mediated Sexual Fantasy (Figure 1). Sexual Scripting and Health Narratives included first-person sexual narratives, protection and contraception content, casual sexual encounter scripts, noncommittal relationship norms, dating platform content, and sexual lip-sync content. Community boundaries were only partly stable: 36.6% of bootstrap samples recovered 4 communities and 51.2% recovered 3. Romantic Relationship Socialization and Mediated Sexual Fantasy items were consistently grouped (item stability, .93 to .98 and .79 to .80), whereas Sexual Scripting and Health Narratives acted as a bridge, its casual-relationship items often grouping with relationship content and its narrative items with sexualized appearance content, as in a 3-community polychoric network (eTable 4). Protection content was consistently grouped with first-person sexual narratives (97% of bootstrap samples) and sexual lip-sync content (90%) but less often with hookup or dating-app content (33% to 39%).

**Figure 1.**
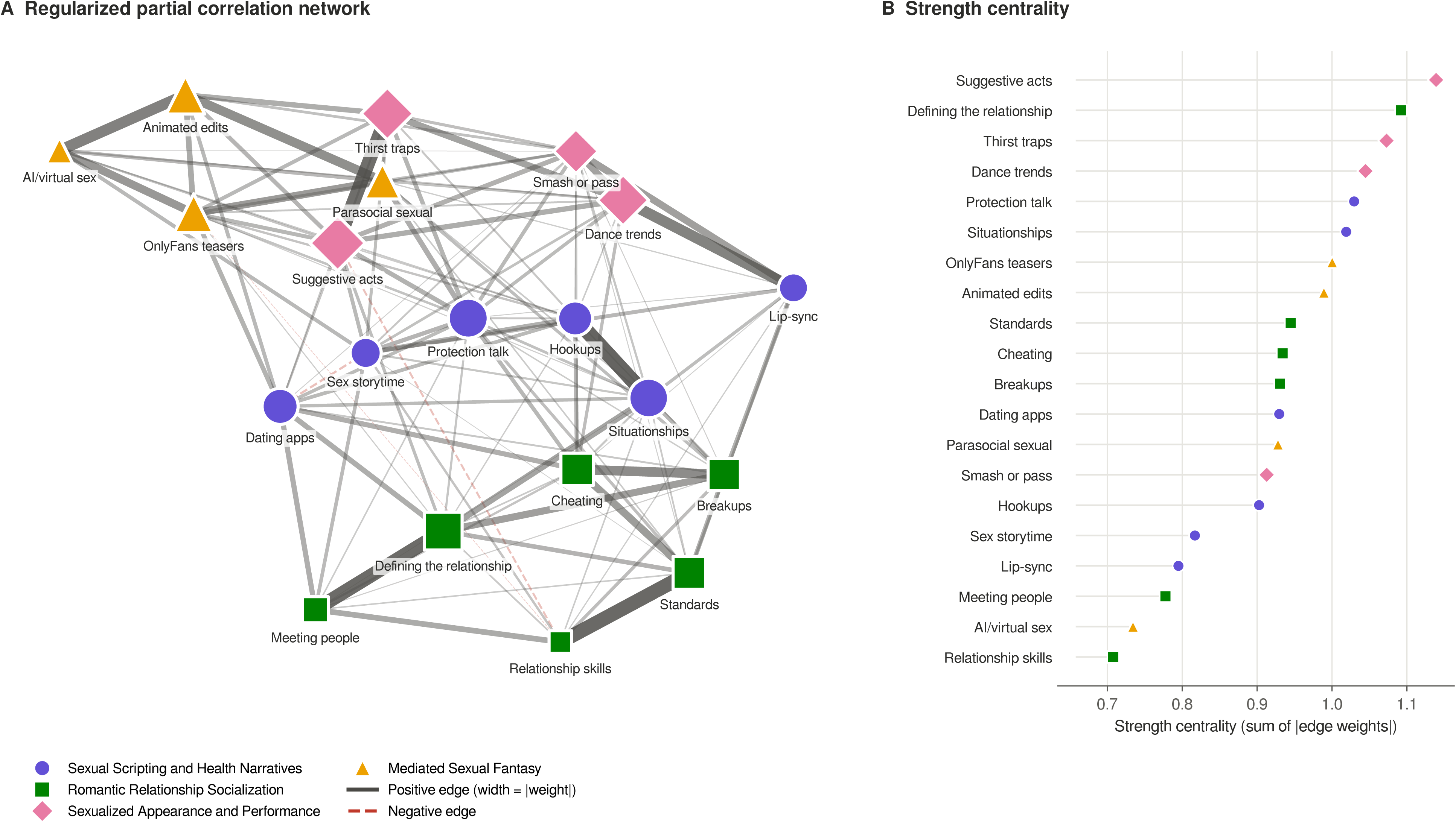
Network of Reported Sexual and Relationship Content Exposure. Panel A shows regularized partial correlations among recalled exposure ratings; node color and shape indicate network-derived communities, node size is proportional to strength centrality, and edge width is proportional to absolute edge weight (dashed red edges are negative). Panel B shows strength centrality. The network included 965 participants with complete valid responses to all 20 items.

Strength centrality reflects how strongly reported exposure to each content category is statistically connected to reported exposure to other categories; higher values indicate stronger overall connections with other exposure ratings, but not more frequent exposure to the content itself. Strength centrality was stable (correlation stability coefficient = 0.75). Suggestive acts, defining the relationship, and thirst traps had the highest strength centrality (Figure 1B).

### Exposure Profiles

A 4-profile solution was selected (eTable 2). BIC improvements diminished after 4 profiles (reductions for 2 to 7 profiles: 2021, 450, 131, 86, 28, 19), and the bootstrap likelihood ratio test, which favored each additional profile through 6 (all P = .01), is known to overextract in large samples. Solutions with 5 or more profiles had lower classification certainty, and the 6-profile solution included a profile of 15 adolescents. For the 4-profile solution, entropy was 0.79, average posterior probabilities ranged from 0.82 to 0.94, and 76.0% of adolescents had a posterior probability of .80 or higher (eTable 3).

The Relationship-Socialization Exposure profile included 284 adolescents (25.9%) and showed moderate exposure to Romantic Relationship Socialization content with low exposure to other communities. The Moderate General Exposure profile included 314 adolescents (28.6%) and showed moderate, relatively balanced exposure across Sexual Scripting and Health Narratives, Romantic Relationship Socialization, and Sexualized Appearance and Performance, with lower Mediated Sexual Fantasy exposure. The High Overall Exposure profile included 299 adolescents (27.3%) and showed frequent exposure across all communities (means, 3.38-3.86), with Sexualized Appearance and Performance content slightly higher than other communities.

The Low-Intensity Relationship Exposure profile included 199 adolescents (18.2%) and showed the lowest overall exposure, relatively concentrated in Romantic Relationship Socialization content (Figure 2).

**Figure 2.**
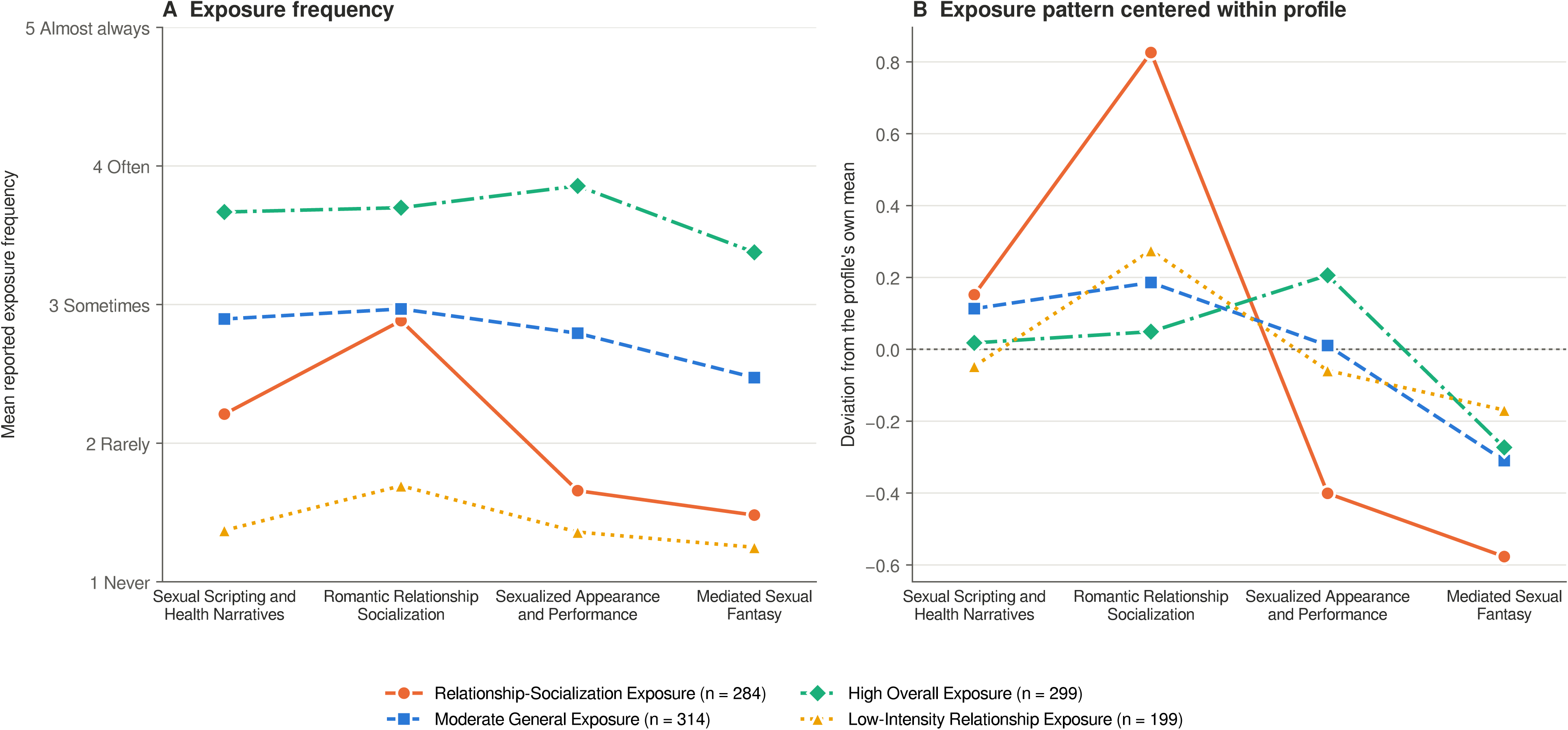
Exposure Frequencies and Within-Profile Patterns. Panel A shows mean recalled exposure frequency for each community by profile. Panel B shows community exposure scores centered on each profile’s overall mean. Latent profile analysis required at least 1 valid item response per community (n = 1096).

Profiles differed by gender, sexual experience, and age, but not sexual minority status (Table 1). Compared with other profiles, the High Overall Exposure profile included more male adolescents and more adolescents who had ever had sex (44.1%) or were currently dating (60.9%), and reported more daily social media time and higher sensation seeking (Table 1). Among adolescents who reported seeing sexual content, 52.4% said it came mostly or always from recommended accounts they did not follow, and 16.0% said it came mostly or always from accounts they followed (eTable 8).

### Profile Associations With Sexual Health Attitudes

Adjusted mean scores for permissive sexual attitudes were highest in the High Overall Exposure profile (3.14; 95% CI, 3.04-3.23), followed by Moderate General Exposure (2.56; 95% CI, 2.47-2.66), Relationship-Socialization Exposure (2.18; 95% CI, 2.08-2.28), and Low-Intensity Relationship Exposure (2.16; 95% CI, 2.04-2.28) (Figure 3A). Compared with Moderate General Exposure, scores were higher in High Overall Exposure (difference, 0.58; 95% CI, 0.40 to 0.75; d = 0.69) and lower in Relationship-Socialization Exposure (difference, −0.38; 95% CI, −0.56 to −0.20) and Low-Intensity Relationship Exposure (difference, −0.40; 95% CI, −0.60 to −0.20; all Tukey-adjusted P < .001).

**Figure 3.**
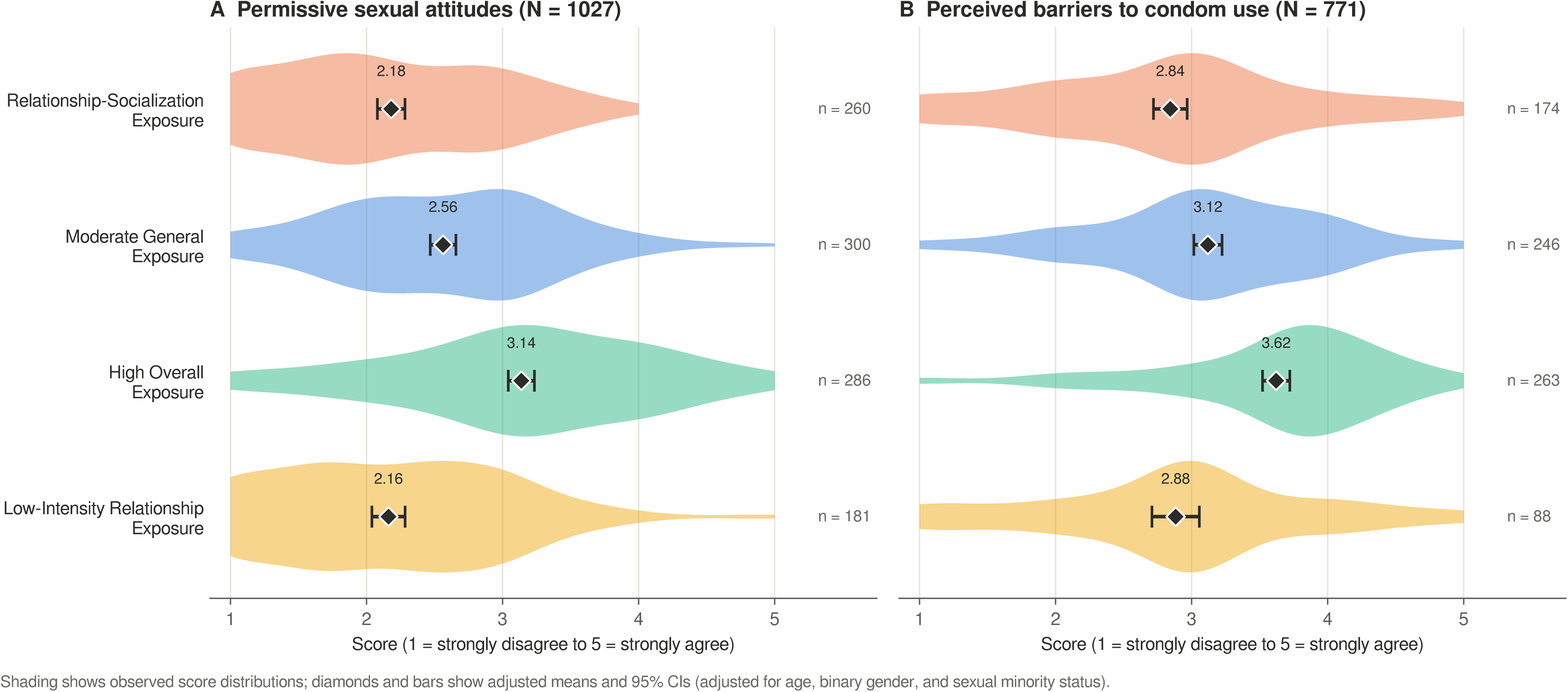
Adjusted Sexual Health Attitudes by Exposure Profile. Shading represents observed score distributions. Diamonds denote adjusted estimated marginal means; error bars indicate pointwise 95% CIs. Models adjusted for age, binary gender, and sexual minority status (permissive sexual attitudes, n = 1027; perceived barriers to condom use, n = 771).

Adjusted mean scores for perceived barriers to condom use were also highest in High Overall Exposure (3.62; 95% CI, 3.52-3.72), followed by Moderate General Exposure (3.12; 95% CI, 3.02-3.22), Low-Intensity Relationship Exposure (2.88; 95% CI, 2.71-3.06), and Relationship-Socialization Exposure (2.84; 95% CI, 2.72-2.97) (Figure 3B). Compared with Moderate General Exposure, scores were higher in High Overall Exposure (difference, 0.50; 95% CI, 0.31 to 0.69; d = 0.61; Tukey-adjusted P < .001) and lower in Relationship-Socialization Exposure (difference, −0.28; 95% CI, −0.49 to −0.07; Tukey-adjusted P = .004). Low-Intensity Relationship Exposure did not differ significantly from Moderate General Exposure (difference, −0.24; 95% CI, −0.50 to 0.03; Tukey-adjusted P = .10).

### Sensitivity Analyses

The difference between the High Overall and Moderate General Exposure profiles was attenuated by approximately one-quarter to one-third but remained significant after additional adjustment for sociodemographic characteristics, religiosity, sensation seeking, and social media time (permissive attitudes, 0.39; 95% CI, 0.20 to 0.58; condom-use barriers, 0.38; 95% CI, 0.17 to 0.59) and after further adjustment for sexual experience and dating status (Table 2). Results were similar with BCH correction, among adolescents with high classification certainty, after excluding possible careless responders (least common in the High Overall Exposure profile: 1.0% vs 4.0% to 5.4%), with categorical gender identity, with complete outcome scales, among sexually experienced adolescents, and when profiles were derived from the 3-community structure (84.6% agreement with primary profiles) (Table 2). Lower scores in the Relationship-Socialization and Low-Intensity Relationship Exposure profiles were robust for permissive attitudes but not consistently for condom-use barriers (eTable 5).

**Table 2.** Adjusted Differences Between the High Overall Exposure and Moderate General Exposure Profiles Across Sensitivity Analyses.

| **Analysis** | **n** | **Permissive attitudes, difference (95% CI)** | **d** | **n** | **Condom-use barriers, difference (95% CI)** | **d** |
|---|---|---|---|---|---|---|
| Primary model | 1027 | 0.58 (0.40 to 0.75) | 0.69 | 771 | 0.50 (0.31 to 0.69) | 0.61 |
| **Additional adjustment** |  |  |  |  |  |  |
| + Sociodemographic characteristics<sup>a</sup> | 1027 | 0.52 (0.33 to 0.71) | 0.63 | 771 | 0.45 (0.25 to 0.65) | 0.54 |
| + Religiosity, sensation seeking, and social media time | 941 | 0.39 (0.20 to 0.58) | 0.50 | 724 | 0.38 (0.17 to 0.59) | 0.47 |
| + Sexual experience and current dating status | 877 | 0.39 (0.20 to 0.58) | 0.50 | 673 | 0.34 (0.12 to 0.56) | 0.42 |
| **Classification uncertainty** |  |  |  |  |  |  |
| BCH-weighted three-step estimation | 1027 | 0.62 (0.40 to 0.84) | 0.77 | 771 | 0.56 (0.35 to 0.76) | 0.68 |
| Posterior probability ≥ .80 only | 783 | 0.65 (0.45 to 0.85) | 0.77 | 608 | 0.58 (0.37 to 0.79) | 0.71 |
| **Measurement and data quality** |  |  |  |  |  |  |
| Profiles derived from 3-community structure<sup>d</sup> | 1029 | 0.56 (0.37 to 0.74) | 0.67 | 772 | 0.51 (0.31 to 0.70) | 0.61 |
| Excluding possible careless responders<sup>b</sup> | 990 | 0.60 (0.42 to 0.78) | 0.72 | 738 | 0.49 (0.30 to 0.69) | 0.60 |
| Gender identity coded categorically<sup>c</sup> | 1060 | 0.56 (0.38 to 0.74) | 0.67 | 787 | 0.50 (0.31 to 0.69) | 0.61 |
| Complete outcome scales only | 1002 | 0.56 (0.38 to 0.74) | 0.68 | 612 | 0.53 (0.34 to 0.72) | 0.70 |
| **Subgroup** |  |  |  |  |  |  |
| Sexually experienced adolescents only | 321 | 0.47 (0.17 to 0.76) | 0.54 | 310 | 0.38 (0.10 to 0.67) | 0.46 |

Differences are adjusted mean differences (High Overall minus Moderate General Exposure) on 1-to-5 scales with Tukey-adjusted simultaneous 95% CIs; all Tukey-adjusted P ≤ .004. d indicates the difference divided by the residual SD. The primary model adjusted for age, binary gender, and sexual minority status; additional covariates were added cumulatively. BCH indicates bias-adjusted three-step estimation. Fit indices for 1 to 7 profiles and all pairwise contrasts are provided in eTables 2 and 5 in the Supplement. <sup>a</sup>Race and ethnicity, school type, parental education, and family financial status. <sup>b</sup>Identical answers to 15 or more consecutive attitude items, identical nonzero answers to all exposure items, or inconsistent reports of never seeing content in 2 or more content blocks (n = 44). <sup>c</sup>Female, male, and nonbinary or another identity; retains nonbinary adolescents excluded from the primary model. <sup>d</sup>Profiles re-estimated from 3 composites (sexual content; romantic, casual, and dating content; mediated sexual fantasy) reflecting the polychoric-network communities.

Each 1-SD increase in overall exposure was associated with higher permissive attitudes (B = 0.39; 95% CI, 0.34-0.44) and perceived condom-use barriers (B = 0.33; 95% CI, 0.27-0.39). Entering the 4 community scores simultaneously modestly improved prediction of permissive attitudes (ΔR^2^ = 0.013; F(3, 1019) = 5.80; P < .001), with unique associations for Sexual Scripting and Health Narratives (B = 0.19), Mediated Sexual Fantasy (B = 0.16), and Sexualized Appearance and Performance (B = 0.13) content but not Romantic Relationship Socialization content (B = −0.05; P = .16). It did not improve prediction of condom-use barriers (ΔR^2^ = 0.005; P = .20). In an exploratory analysis holding exposure to all other content constant, more frequent exposure to protection and contraception content was associated with greater perceived condom-use barriers (per SD, B = 0.09; 95% CI, 0.01-0.17) and more permissive attitudes (B = 0.13; 95% CI, 0.06-0.21), including after adjustment for sexual experience (eTable 6).

Overall, 278 adolescents (24.7%) answered “not sure/not applicable” to all 4 condom-barrier items, including 35.4% of those who had not had sex and 3.7% of those who had. This response was most common in the Low-Intensity Relationship Exposure profile (49.2%) and least common in the High Overall Exposure profile (8.4%) (adjusted odds ratios vs Moderate General Exposure, 4.54 [95% CI, 2.97-6.94] and 0.38 [95% CI, 0.22-0.65]), and differences persisted after adjustment for sexual experience (eTable 7).

## Discussion

In this quota-based online sample of US adolescents, reported exposure to sexual and relationship content on social media was patterned, and profiles differed in both overall exposure frequency and the types of content adolescents encountered. Adolescents in the High Overall Exposure profile reported the most permissive sexual attitudes and the greatest perceived condom-use barriers. These associations were robust to data-quality exclusions and correction for classification error and were attenuated, but not eliminated, by adjustment for sensation seeking, religiosity, social media time, and sociodemographic characteristics.

One methodological contribution is a content-specific exposure measure developed from existing research, structured review of contemporary content, and adolescent input [33], capturing platform-native content such as situationships, thirst traps, and interactive sexual trends. Suggestive acts, thirst traps, and defining the relationship had the highest strength centrality. Centrality in a cross-sectional network does not indicate a temporal or causal role; whether relationship-status content serves as an entry point into broader sexual content, for example through engagement-based recommendations, requires longitudinal or platform-level data. Notably, about half of adolescents who saw sexual content attributed it mainly to recommended accounts they did not follow, consistent with often incidental exposure.

Protection and contraception content was consistently grouped with first-person sexual narratives and sexual lip-sync content, rather than with relationship content or as a separate health-information community. On social media, adolescents therefore appear to encounter protection mainly alongside other sexual content, possibly through creators’ storytimes rather than health sources. This placement cuts both ways. Peer-voiced narratives can make protection relatable, but they may also portray condoms as awkward, unnecessary, or at odds with intimacy. Consistent with the latter possibility, more frequent exposure to protection content, holding other exposure constant, was associated with greater, not fewer, perceived condom-use barriers; alternatively, adolescents with concerns about condoms may seek out such content. Because this association was small, cross-sectional, and based on a single item that did not capture message framing, exposure to protection-related topics should not be equated with exposure to supportive sexual health messages. Content analyses of how protection is framed within these narratives are needed.

At the person level, both the amount and the nature of exposure mattered, although not equally. The High Overall Exposure profile had the most permissive attitudes and greatest perceived condom-use barriers, and overall exposure frequency accounted for most of these associations. Content type nonetheless shaped them. The Relationship-Socialization profile reported more exposure than the Low-Intensity Relationship profile, mostly to relationship content, yet the 2 profiles did not differ on either outcome (eTable 5); the Moderate General profile’s additional sexual content, in contrast, accompanied more permissive attitudes. In continuous models, sexual narratives, mediated sexual fantasy, and sexualized appearance content, but not relationship content, were uniquely associated with permissive attitudes, whereas content type added little for condom-use barriers. Thus, exposure to relationship-focused content was not associated with these attitudes, whereas exposure to sexual content was. From sexual script and social cognitive perspectives, repeated and sometimes incidental exposure may provide models of sexual behavior, relationship expectations, and protection that inform perceptions of what is typical or acceptable [12,13]. Selective exposure is an equally plausible explanation [34–36]: adolescents who already hold more permissive attitudes, are higher in sensation seeking, or are sexually experienced may seek out and engage with sexual content, which recommendation systems could then amplify. Consistent with this, the High Overall Exposure profile reported higher sensation seeking and more sexual experience, and adjusting for these factors attenuated the associations. Longitudinal research is needed to disentangle these processes.

One-quarter of adolescents answered “not sure/not applicable” to every condom-barrier item, most often those without sexual experience and those with the least exposure. For these adolescents, condom-related beliefs may still be forming, which may make early, accurate counseling particularly valuable.

These findings have implications for adolescent sexual health counseling and media literacy. Approaches focused on whether adolescents encounter accurate sexual health information may overlook how much sexual content they encounter and how protection is portrayed within it. Clinicians and educators could encourage adolescents to reflect on the content they see, including content recommended to them, and to distinguish reliable information from messages that portray protection as undesirable. Whether such approaches improve sexual health attitudes or behavior warrants evaluation.

### Limitations

Several limitations warrant consideration. The cross-sectional design cannot establish the temporal direction of associations, and residual confounding is possible. Exposure was assessed through adolescents’ recall of typical content frequencies rather than direct observation and did not capture message framing; the new measure requires further validation, and its community boundaries, particularly for casual-relationship content, were not fully stable, although profile associations were similar under an alternative 3-community structure. The quota-based, unweighted online sample limits generalizability, and the primary models’ binary gender covariate excluded adolescents with other identities, although results were similar when gender identity was coded categorically. Outcomes were attitudes, not behavior or sexually transmitted infections; the permissive attitudes composite was not a previously validated scale and included an item on condomless sex, and many adolescents without sexual experience could not rate condom-use barriers.

Future studies should validate adolescent-informed exposure measures against observed social media content and use longitudinal designs and more representative samples to examine how exposure patterns and sexual health attitudes change over time.

### Conclusions

Adolescents’ reported exposure to sexual and relationship content on social media followed distinct patterns, and higher overall exposure was associated with more permissive sexual attitudes and greater perceived barriers to condom use. These findings highlight everyday social media use as an important context for adolescent sexual socialization and suggest that sexual health counseling may benefit from greater attention to the range of sexual and relationship messages adolescents encounter online. Longitudinal research incorporating direct observation of social media exposure is needed to examine how these patterns develop and relate to changes in sexual health beliefs.

## Supporting information

Supplementary Online Content

## Data Availability

All data produced in the present study are available upon reasonable request to the authors

## Notes

### Competing Interest Statement

The Digital Wellness Lab has received unrestricted research funding from multiple organizations. These organizations were not involved in the design, conduct, analysis, or reporting of this study. A list of supporting organizations is available at https://digitalwellnesslab.org/supporters/.

### Author Declarations

The Institutional Review Board of Boston Children's Hospital gave ethical approval for this work.

