## Supplementary Online Content for "Sexual and Relationship Content Exposure on Social Media and Adolescent Sexual Health Attitudes"

eTable 1. Sexual and Relationship Content Exposure Items and Descriptive Statistics

eTable 2. Latent Profile Model Fit Indices for 1 to 7 Profiles

eTable 3. Community Exposure Means and Average Posterior Classification Probabilities by Profile

eTable 4. Network Robustness: Centrality, Community Stability, and Polychoric Sensitivity Network

eTable 5. Adjusted Pairwise Profile Contrasts Across Primary and Sensitivity Analyses

eTable 6. Associations of Overall Exposure and Community-Specific Exposure With Outcomes

eTable 7. "Not Sure/Not Applicable" Responses to Condom-Barrier Items by Profile

eTable 8. Reported Source of Sexual and Relationship Content

eFigure 1. Bootstrap Community Co-Assignment of Exposure Items

eFigure 2. Stability of Strength Centrality Under Case-Dropping Bootstrap

*This supplementary material has been provided by the authors to give readers additional information about their work.*

### eMethods

#### Data quality indicators

Two embedded attention-check items were used during data collection, and responses failing either check were excluded before the analytic data set was assembled. Because attention checks detect only some forms of inattentive responding, we examined three additional indicators of possible careless responding (Meade & Craig, 2012): (1) a longstring index of 15 or more identical consecutive responses across 18 attitude items that differed in item direction (e.g., "I need to be committed to a person to have sex with them" and "One-night stands can be very enjoyable"; "not sure/not applicable" responses were treated as breaks) ($n = 30$); (2) identical nonzero responses (rarely through almost always) to all 20 exposure items ($n = 5$), excluding adolescents who answered "never" to all items, which is a plausible response pattern; and (3) in 2 or more of the 5 content blocks, reporting "I haven't seen this type of content" on the content-source item after rating at least 1 item in that block as seen often or almost always ($n = 14$). In total, 44 adolescents (3.9%) met at least 1 criterion. A broader definition that also flagged adolescents who strongly agreed both that they needed commitment to have sex and that one-night stands can be very enjoyable flagged 72 adolescents. Outcome models were re-estimated excluding flagged adolescents, retaining their original profile assignments, and after re-estimating the latent profile model in the remaining sample (97.8% of adolescents retained the same profile assignment). No duplicate IP addresses or session identifiers were present.

#### Latent profile estimation

Latent profile models were Gaussian mixtures with equal within-profile variances and zero within-profile covariances (mclust model EEI), estimated with 100 random starts (k-means and random-partition initializations) and a relative log-likelihood convergence tolerance of 10^-10. All random starts converged to the same 4-profile solution. Because the default mclust convergence tolerance (10^-5) can stop the expectation-maximization algorithm before full convergence when profiles overlap, we recommend specifying a strict tolerance (e.g., emControl(tol = c(1e-10, sqrt(.Machine$double.eps)))) for reproduction. The bootstrap likelihood ratio test compared k - 1 and k profiles using 100 parametric bootstrap samples generated from the k - 1 profile model, each fitted with multiple random starts.

#### Correction for classification error

Assigning adolescents to their most likely profile and treating assignments as known can bias associations with outcomes. We therefore used the bias-adjusted three-step BCH approach (Vermunt, 2010; Bakk & Vermunt, 2016). The classification error matrix D, with elements D(t, s) = P(assigned profile = s | true profile = t), was estimated from the posterior probabilities. Each adolescent contributed 1 record per latent profile t, weighted by the element of D^-1 corresponding to the adolescent's assigned profile and profile t. Weighted least squares models including the same covariates as the primary analysis were estimated, with cluster-robust standard errors accounting for multiple records per adolescent.

#### Network stability

Strength-centrality stability was assessed with 1,000 case-dropping bootstrap samples that dropped 5% to 75% of participants, re-estimating the EBICglasso network in each sample; the correlation stability coefficient is the maximum proportion of cases that can be dropped while retaining, with 95% probability, a correlation of at least 0.70 with the original centrality indices. Community stability was assessed with 500 nonparametric bootstrap samples (resampling participants with replacement), re-estimating the network and Walktrap communities in each sample. We recorded the number of communities, the proportion of samples in which each pair of items was assigned to the same community (eFigure 1), and item stability (proportion of samples in which an item was assigned to the bootstrap community best matching its empirical community). A sensitivity network used polychoric correlations estimated by maximum likelihood.

**Covariate coding**

Race and ethnicity were categorized as Hispanic or Latino (any race), non-Hispanic White, Black, or Asian (single race), another race or multiple races, and not reported. School type was categorized as traditional public; charter, magnet, or technical or vocational; and private, homeschool, or other. Parental education (high school or less, some college or associate degree, bachelor's degree or higher, unknown) and perceived family financial status (below average, average, above average, not reported) were categorical. Religiosity was coded 0 for adolescents reporting no religious affiliation and 1 to 5 for closeness of identification with one's religion. Sensation seeking was the mean of the 8-item Brief Sensation Seeking Scale (Hoyle et al., 2002). Social media time was the sum of category midpoints for typical daily time on YouTube, Instagram, and TikTok (0 for nonusers). Current dating status was coded 1 for adolescents currently in or "talking to" 1 or more romantic partners.

#### eTable 1. Sexual and Relationship Content Exposure Items and Descriptive Statistics

| Item | Survey block | Mean (SD) | Often or almost always, % | Never, % | Prefer not to answer / don't remember, No. |
|---|---|---|---|---|---|
| **Sexual Scripting and Health Narratives** | | | | | |
| Personal sexual story-times or advice (verbal discussion, not visually explicit) | Sexual content and behaviors | 2.38 (1.14) | 17.1 | 27.3 | 31 |
| Protection and contraception talk (condoms, birth control, plan B) | Sexual content and behaviors | 2.43 (1.24) | 21.9 | 29.8 | 26 |
| Lip-syncing videos to explicit lyrics | Sexual content and behaviors | 3.08 (1.38) | 42.0 | 19.0 | 29 |
| Dating apps: posts that discusses using dating apps (e.g., Tinder, Bumble, Hinge, Grindr, etc.) | Relationship initiation | 2.42 (1.30) | 24.8 | 34.2 | 26 |
| Hook-up culture (one-night stands, body count jokes, walk-of-shame memes) | Casual, noncommittal encounters | 2.77 (1.30) | 29.7 | 22.9 | 35 |
| Ongoing casual situations (friends with benefits, situationships, no labels) | Casual, noncommittal encounters | 2.83 (1.28) | 32.2 | 20.4 | 33 |
| **Romantic Relationship Socialization** | | | | | |
| How to meet people, start talking, or plan a first date | Relationship initiation | 2.56 (1.12) | 19.0 | 20.4 | 35 |
| Defining the relationship/talking stage (e.g., posts about deciding what you are, exclusivity conversations, situationsh… | Relationship initiation | 2.76 (1.21) | 30.4 | 18.9 | 43 |
| Healthy-relationship skills: advice on communication, boundary-setting, conflict resolution, trust building | Ongoing relationships and breakups | 2.95 (1.12) | 30.2 | 11.9 | 47 |
| Standards & expectations: list of bare minimums, green/red flags, or gestures partners should do | Ongoing relationships and breakups | 3.22 (1.12) | 41.4 | 8.3 | 38 |
| Cheating accusations, jealousy, or checking your partner /loyalty tests | Ongoing relationships and breakups | 2.98 (1.24) | 38.4 | 15.4 | 37 |
| Break-up experience and recovery: story-time about break-ups, heart break, glow up arcs | Ongoing relationships and breakups | 3.02 (1.26) | 35.1 | 15.0 | 37 |
| **Sexualized Appearance and Performance** | | | | | |
| Near-nudity/thirst traps (often features more explicit or suggestive visuals/display of one's body) | Sexual content and behaviors | 2.54 (1.33) | 24.5 | 28.7 | 30 |
| Sexually suggestive gestures/simulated sexual acts (dry-humping, thrusting, oral gestures) | Sexual content and behaviors | 2.31 (1.27) | 19.6 | 36.9 | 31 |
| Sexualized dance trends (twerking, slow body rolls) | Sexual content and behaviors | 2.77 (1.37) | 32.4 | 24.2 | 28 |
| Interactive sexually themed games/challenges/filters: smash or pass, rate my body | Sexual content and behaviors | 2.49 (1.26) | 22.4 | 28.8 | 30 |
| **Mediated Sexual Fantasy** | | | | | |
| AI-generated or virtual influencer dating/sexual content | Animated, edited, or promotional content | 2.09 (1.16) | 13.1 | 40.5 | 28 |
| Animated sexual edits (e.g., anime, cartoons) | Animated, edited, or promotional content | 2.26 (1.22) | 18.7 | 37.8 | 35 |
| Paid-content teasers (e.g., OnlyFans link in bio) | Animated, edited, or promotional content | 2.25 (1.28) | 20.1 | 40.5 | 27 |
| Parasocial romantic or sexual content: posts or edits that portrays a public figure (celebrity or influencer) or fiction… | Animated, edited, or promotional content | 2.33 (1.29) | 20.6 | 36.8 | 33 |

Participants were asked how often they see each type of content during their usual social media use (1 = never, 2 = rarely, 3 = sometimes, 4 = often, 5 = almost always, 6 = prefer not to answer/I don't remember; 6 treated as missing). Items are grouped by network-derived community. Cronbach α: all 20 items, .94; communities, .82-.86. After each survey block, participants were asked where such content usually came from (eTable 8). Item wording is reproduced from the survey data file; wording marked … was truncated in the data file and appears in full in the survey instrument.

#### eTable 2. Latent Profile Model Fit Indices for 1 to 7 Profiles

| Profiles | Log-likelihood | AIC | BIC | SABIC | Entropy | Minimum average posterior probability | Smallest profile, No. (%) | BLRT P (k - 1 vs k) |
|---|---|---|---|---|---|---|---|---|
| 1 | -6210.7 | 12437.3 | 12477.3 | 12451.9 | — | — | — | — |
| 2 | -5182.8 | 10391.5 | 10456.5 | 10415.2 | 0.881 | 0.965 | 521 (47.5) | .010 |
| 3 | -4940.4 | 9916.9 | 10006.9 | 9949.7 | 0.832 | 0.885 | 340 (31.0) | .010 |
| 4 | -4857.4 | 9760.7 | 9875.7 | 9802.7 | 0.789 | 0.820 | 199 (18.2) | .010 |
| 5 | -4796.9 | 9649.8 | 9789.8 | 9700.9 | 0.795 | 0.776 | 120 (10.9) | .010 |
| 6 | -4765.3 | 9596.6 | 9761.6 | 9656.8 | 0.825 | 0.788 | 15 (1.4) | .010 |
| 7 | -4738.5 | 9553.0 | 9743.0 | 9622.3 | 0.838 | 0.785 | 15 (1.4) | — |

n = 1096. Models assumed equal within-profile variances and zero covariances and were estimated with 100 random starts. AIC indicates Akaike information criterion; BIC, Bayesian information criterion; BLRT, bootstrap likelihood ratio test (100 bootstrap samples; the smallest attainable P value is .01); SABIC, sample size-adjusted BIC. The 4-profile solution was selected because BIC improvements diminished after 4 profiles, solutions with 5 or more profiles had lower classification certainty, and the 6- and 7-profile solutions included a profile of 15 adolescents.

#### eTable 3. Community Exposure Means and Average Posterior Classification Probabilities by Profile

| Profile | No. | Sexual Scripting and Health Narratives | Romantic Relationship Socialization | Sexualized Appearance and Performance | Mediated Sexual Fantasy | P(Relationship-Socialization) | P(Moderate General) | P(High Overall) | P(Low-Intensity Relationship) |
|---|---|---|---|---|---|---|---|---|---|
| Relationship-Socialization | 284 | 2.21 (0.47) | 2.88 (0.55) | 1.66 (0.47) | 1.48 (0.51) | 0.82 | 0.08 | 0.00 | 0.10 |
| Moderate General | 314 | 2.90 (0.52) | 2.97 (0.62) | 2.79 (0.55) | 2.47 (0.70) | 0.07 | 0.86 | 0.06 | 0.00 |
| High Overall | 299 | 3.67 (0.42) | 3.70 (0.50) | 3.86 (0.51) | 3.38 (0.64) | 0.00 | 0.06 | 0.94 | 0.00 |
| Low-Intensity Relationship | 199 | 1.37 (0.37) | 1.69 (0.56) | 1.36 (0.47) | 1.25 (0.43) | 0.10 | 0.00 | 0.00 | 0.89 |

Community means are on the original 1-to-5 frequency scale (SD in parentheses). Posterior columns show the mean posterior probability of membership in each profile among adolescents assigned to the row profile; diagonal values are average posterior probabilities for the assigned profile. Overall, 76.0% of adolescents had a maximum posterior probability of .80 or higher.

#### eTable 4. Network Robustness: Centrality, Community Stability, and Polychoric Sensitivity Network

| Item | Community | Strength | Item stability | Co-assigned with protection talk | Strength (polychoric) | Polychoric community |
|---|---|---|---|---|---|---|
| Suggestive acts | Sexualized Appearance and Performance | 1.139 | 0.64 | 0.45 | 1.173 | 2 |
| Defining the relationship | Romantic Relationship Socialization | 1.092 | 0.97 | 0.02 | 1.093 | 3 |
| Thirst traps | Sexualized Appearance and Performance | 1.073 | 0.63 | 0.43 | 1.064 | 2 |
| Dance trends | Sexualized Appearance and Performance | 1.045 | 0.75 | 0.70 | 1.043 | 2 |
| Protection talk | Sexual Scripting and Health Narratives | 1.030 | 0.53 | — | 1.032 | 2 |
| Situationships | Sexual Scripting and Health Narratives | 1.019 | 0.47 | 0.28 | 1.022 | 3 |
| OnlyFans teasers | Mediated Sexual Fantasy | 1.000 | 0.80 | 0.08 | 1.010 | 1 |

| Item | Community | Strength | Item stability | Co-assigned with protection talk | Strength (polychoric) | Polychoric community |
|---|---|---|---|---|---|---|
| Animated edits | Mediated Sexual Fantasy | 0.989 | 0.80 | 0.08 | 0.998 | 1 |
| Standards | Romantic Relationship Socialization | 0.945 | 0.98 | 0.02 | 0.945 | 3 |
| Cheating | Romantic Relationship Socialization | 0.934 | 0.93 | 0.05 | 0.917 | 3 |
| Breakups | Romantic Relationship Socialization | 0.931 | 0.96 | 0.03 | 0.947 | 3 |
| Dating apps | Sexual Scripting and Health Narratives | 0.929 | 0.59 | 0.39 | 0.907 | 3 |
| Parasocial sexual | Mediated Sexual Fantasy | 0.928 | 0.79 | 0.09 | 0.945 | 1 |
| Smash or pass | Sexualized Appearance and Performance | 0.913 | 0.75 | 0.71 | 0.922 | 2 |
| Hookups | Sexual Scripting and Health Narratives | 0.903 | 0.53 | 0.33 | 0.904 | 3 |
| Sex storytime | Sexual Scripting and Health Narratives | 0.817 | 0.55 | 0.97 | 0.805 | 2 |
| Lip-sync | Sexual Scripting and Health Narratives | 0.795 | 0.47 | 0.90 | 0.795 | 2 |
| Meeting people | Romantic Relationship Socialization | 0.778 | 0.97 | 0.02 | 0.806 | 3 |
| AI/virtual sex | Mediated Sexual Fantasy | 0.734 | 0.80 | 0.08 | 0.807 | 1 |
| Relationship skills | Romantic Relationship Socialization | 0.708 | 0.98 | 0.02 | 0.701 | 3 |

Primary network: n = 965; 115 of 190 possible edges were nonzero; modularity = 0.28. Correlation stability coefficients (case-dropping bootstrap, 1,000 samples): strength, 0.75; closeness, 0.21; betweenness, 0.05. Number of communities across 500 nonparametric bootstrap samples: 2 communities, 10.2%, 3 communities, 51.2%, 4 communities, 36.6%, 5 communities, 2.0%. Item stability is the proportion of bootstrap samples in which an item was assigned to the community best matching its empirical community. Polychoric network: 3 communities (modularity = 0.35); strength centrality correlated r = 0.98 and edge weights correlated r = 1.00 with the primary network. Polychoric community numbers are arbitrary labels.

#### eTable 5. Adjusted Pairwise Profile Contrasts Across Primary and Sensitivity Analyses

*Permissive sexual attitudes*

| Analysis | n | HO - MG | MG - RS | LI - MG | HO - RS | LI - RS | LI - HO |
|---|---|---|---|---|---|---|---|
| Primary (age, gender, sexual minority) | 1027 | 0.58 (<.001) | 0.38 (<.001) | -0.40 (<.001) | 0.96 (<.001) | -0.02 (.99) | -0.98 (<.001) |
| + race/ethnicity, school type, parent education, family finances | 1027 | 0.52 (<.001) | 0.37 (<.001) | -0.40 (<.001) | 0.89 (<.001) | -0.03 (.98) | -0.92 (<.001) |
| + sociodemographics, religiosity, sensation seeking, social media time | 941 | 0.39 (<.001) | 0.26 (.001) | -0.29 (.002) | 0.66 (<.001) | -0.03 (.99) | -0.68 (<.001) |
| + all above, ever had sex, currently dating | 877 | 0.39 (<.001) | 0.26 (.003) | -0.28 (.006) | 0.64 (<.001) | -0.03 (.99) | -0.67 (<.001) |
| + ever had sex (original sensitivity analysis) | 960 | 0.57 (<.001) | 0.36 (<.001) | -0.37 (<.001) | 0.93 (<.001) | -0.01 (1.00) | -0.94 (<.001) |
| Gender identity coded categorically (retains nonbinary youth) | 1060 | 0.56 (<.001) | 0.38 (<.001) | -0.38 (<.001) | 0.94 (<.001) | -0.01 (1.00) | -0.94 (<.001) |
| Excluding flagged careless responders | 990 | 0.60 (<.001) | 0.41 (<.001) | -0.41 (<.001) | 1.00 (<.001) | -0.00 (1.00) | -1.00 (<.001) |
| Excluding careless responders (broad definition) | 966 | 0.54 (<.001) | 0.40 (<.001) | -0.41 (<.001) | 0.94 (<.001) | -0.01 (1.00) | -0.96 (<.001) |
| Restricted to posterior probability >= .80 | 783 | 0.65 (<.001) | 0.35 (<.001) | -0.36 (<.001) | 0.99 (<.001) | -0.02 (1.00) | -1.01 (<.001) |
| Complete 4-item scores only | 1002 | 0.56 (<.001) | 0.38 (<.001) | -0.41 (<.001) | 0.94 (<.001) | -0.03 (.98) | -0.97 (<.001) |
| Sexually experienced adolescents only | 321 | 0.47 (<.001) | 0.45 (.008) | -0.09 (.96) | 0.92 (<.001) | 0.36 (.28) | -0.56 (.01) |
| BCH-corrected for classification error | 1027 | 0.62 (<.001) | 0.41 (<.001) | -0.39 (<.001) | 1.03 (<.001) | 0.02 (1.00) | -1.01 (<.001) |
| LPA re-estimated without careless responders | 990 | 0.62 (<.001) | 0.39 (<.001) | -0.38 (<.001) | 1.01 (<.001) | 0.01 (1.00) | -1.01 (<.001) |
| Profiles derived from 3-community structure | 1029 | 0.56 (<.001) | 0.25 (.004) | -0.50 (<.001) | 0.81 (<.001) | -0.25 (.007) | -1.06 (<.001) |

*Perceived barriers to condom use*

| Analysis | n | HO - MG | MG - RS | LI - MG | HO - RS | LI - RS | LI - HO |
|---|---|---|---|---|---|---|---|
| Primary (age, gender, sexual minority) | 771 | 0.50 (<.001) | 0.28 (.004) | -0.24 (.10) | 0.78 (<.001) | 0.04 (.98) | -0.74 (<.001) |
| + race/ethnicity, school type, parent education, family finances | 771 | 0.45 (<.001) | 0.29 (.003) | -0.24 (.11) | 0.74 (<.001) | 0.05 (.96) | -0.69 (<.001) |
| + sociodemographics, religiosity, sensation seeking, social media time | 724 | 0.38 (<.001) | 0.24 (.03) | -0.12 (.68) | 0.62 (<.001) | 0.12 (.74) | -0.50 (<.001) |
| + all above, ever had sex, currently dating | 673 | 0.34 (<.001) | 0.23 (.05) | -0.14 (.62) | 0.57 (<.001) | 0.09 (.88) | -0.48 (<.001) |
| + ever had sex (original sensitivity analysis) | 718 | 0.48 (<.001) | 0.27 (.009) | -0.26 (.07) | 0.75 (<.001) | 0.01 (1.00) | -0.74 (<.001) |
| Gender identity coded categorically (retains nonbinary youth) | 787 | 0.50 (<.001) | 0.27 (.005) | -0.25 (.08) | 0.77 (<.001) | 0.02 (1.00) | -0.75 (<.001) |
| Excluding flagged careless responders | 738 | 0.49 (<.001) | 0.32 (<.001) | -0.26 (.08) | 0.82 (<.001) | 0.07 (.93) | -0.75 (<.001) |
| Excluding careless responders (broad definition) | 715 | 0.48 (<.001) | 0.32 (.001) | -0.25 (.09) | 0.80 (<.001) | 0.07 (.94) | -0.73 (<.001) |
| Restricted to posterior probability >= .80 | 608 | 0.58 (<.001) | 0.22 (.11) | -0.27 (.09) | 0.80 (<.001) | -0.06 (.97) | -0.86 (<.001) |
| Barrier score requires >= 3 valid items | 674 | 0.53 (<.001) | 0.21 (.06) | -0.30 (.03) | 0.74 (<.001) | -0.09 (.85) | -0.83 (<.001) |
| Complete 4-item scores only | 612 | 0.53 (<.001) | 0.20 (.10) | -0.34 (.01) | 0.73 (<.001) | -0.14 (.62) | -0.87 (<.001) |
| Sexually experienced adolescents only | 310 | 0.38 (.004) | 0.06 (.97) | -0.36 (.23) | 0.44 (.005) | -0.30 (.46) | -0.75 (<.001) |
| Adolescents without sexual experience only | 408 | 0.59 (<.001) | 0.39 (.003) | -0.21 (.38) | 0.98 (<.001) | 0.18 (.56) | -0.81 (<.001) |
| BCH-corrected for classification error | 771 | 0.56 (<.001) | 0.29 (.05) | -0.20 (.47) | 0.85 (<.001) | 0.09 (.95) | -0.75 (<.001) |
| LPA re-estimated without careless responders | 738 | 0.50 (<.001) | 0.28 (.007) | -0.28 (.05) | 0.78 (<.001) | -0.00 (1.00) | -0.78 (<.001) |
| Profiles derived from 3-community structure | 772 | 0.51 (<.001) | 0.21 (.06) | -0.30 (.007) | 0.72 (<.001) | -0.08 (.82) | -0.80 (<.001) |

Values are adjusted mean differences (Tukey-adjusted P). HO indicates High Overall Exposure; LI, Low-Intensity Relationship Exposure; MG, Moderate General Exposure; RS, Relationship-Socialization Exposure. Covariate sets are cumulative; see Table 2 for definitions.

#### eTable 6. Associations of Overall Exposure and Community-Specific Exposure With Outcomes

| Predictor | B (95% CI) | P value | $R^2$ |
|---|---|---|---|
| **Permissive sexual attitudes** | | | |
| Model A: overall exposure (per SD) | 0.39 (0.34 to 0.44) | <.001 | 0.209 |
| Model B: Sexual Scripting and Health Narratives (per SD) | 0.19 (0.10 to 0.29) | <.001 | 0.222 |
| Model B: Romantic Relationship Socialization (per SD) | -0.05 (-0.13 to 0.02) | .16 | |
| Model B: Sexualized Appearance and Performance (per SD) | 0.13 (0.04 to 0.22) | .005 | |
| Model B: Mediated Sexual Fantasy (per SD) | 0.16 (0.08 to 0.24) | <.001 | |
| Model B vs A | $\Delta R^2$ = 0.013; F(3, 1019) = 5.80 | <.001 | |
| Profiles added to Model A | $\Delta R^2$ = 0.016; F(3, 1018) = 6.96 | <.001 | 0.225 |
| **Perceived barriers to condom use** | | | |
| Model A: overall exposure (per SD) | 0.33 (0.27 to 0.39) | <.001 | 0.201 |
| Model B: Sexual Scripting and Health Narratives (per SD) | 0.03 (-0.08 to 0.15) | .60 | 0.206 |
| Model B: Romantic Relationship Socialization (per SD) | 0.05 (-0.03 to 0.14) | .23 | |
| Model B: Sexualized Appearance and Performance (per SD) | 0.11 (0.01 to 0.22) | .03 | |
| Model B: Mediated Sexual Fantasy (per SD) | 0.18 (0.09 to 0.27) | <.001 | |
| Model B vs A | $\Delta R^2$ = 0.005; F(3, 763) = 1.53 | .20 | |
| Profiles added to Model A | $\Delta R^2$ = 0.013; F(3, 762) = 4.36 | .005 | 0.215 |
| **Exploratory: protection and contraception content (per SD), holding other exposure constant** | | | |
| Permissive sexual attitudes: primary covariates (n = 1025) | 0.13 (0.06 to 0.21) | <.001 | |
| Permissive sexual attitudes: + ever had sex (n = 959) | 0.14 (0.06 to 0.21) | <.001 | |
| Permissive sexual attitudes: + ever had sex, sensation seeking, social media time (n = 908) | 0.12 (0.04 to 0.19) | .002 | |
| Perceived barriers to condom use: primary covariates (n = 770) | 0.09 (0.01 to 0.17) | .02 | |
| Perceived barriers to condom use: + ever had sex (n = 718) | 0.12 (0.03 to 0.20) | .007 | |
| Perceived barriers to condom use: + ever had sex, sensation seeking, social media time (n = 691) | 0.10 (0.02 to 0.19) | .01 | |

All models adjusted for age, binary gender, and sexual minority status (permissive attitudes, n = 1027; condom-use barriers, n = 771). Overall exposure is the mean of the 4 community scores. In model B, the 4 standardized community scores were entered simultaneously (maximum variance inflation factor, 3.5). The final row tests whether profile membership explains variance beyond overall exposure. The exploratory protection models regress each outcome on standardized exposure to protection and contraception content and the standardized mean of the other 19 items.

#### eTable 7. "Not Sure/Not Applicable" Responses to Condom-Barrier Items by Profile

| Profile | No. | All 4 items not sure, No. (%) | Any item not sure, % | All 4, % (no sexual experience) | All 4, % (sexually experienced) | OR (95% CI), model 1 | OR (95% CI), model 2 |
|---|---|---|---|---|---|---|---|
| Relationship-Socialization | 284 | 83 (29.2) | 47.9 | 39.6 | 1.5 | 1.84 (1.23-2.75) | 1.58 (1.01-2.45) |
| Moderate General | 314 | 61 (19.4) | 38.9 | 29.2 | 4.5 | 1 [Reference] | 1 [Reference] |
| High Overall | 299 | 25 (8.4) | 18.4 | 13.7 | 2.3 | 0.38 (0.22-0.65) | 0.41 (0.23-0.73) |

| Profile | No. | All 4 items not sure, No. (%) | Any item not sure, % | All 4, % (no sexual experience) | All 4, % (sexually experienced) | OR (95% CI), model 1 | OR (95% CI), model 2 |
|---|---|---|---|---|---|---|---|
| Low-Intensity Relationship | 199 | 98 (49.2) | 63.8 | 58.3 | 12.9 | 4.54 (2.97-6.94) | 3.95 (2.48-6.31) |

Overall, 278 of 1124 adolescents (24.7%) answered "not sure/not applicable" to all 4 condom-barrier items and therefore had no barrier score. Odds ratios are from logistic regression of answering "not sure/not applicable" to all 4 items; model 1 adjusted for age, binary gender, and sexual minority status; model 2 additionally adjusted for ever having had sex.

#### eTable 8. Reported Source of Sexual and Relationship Content

| Content block | No. reporting exposure | Mostly or always from followed accounts, % | About half and half, % | Mostly or always recommended (not followed), % | Not sure or prefer not to answer, % |
|---|---|---|---|---|---|
| Relationship initiation | 881 | 20.7 | 25.0 | 45.6 | 8.7 |
| Ongoing relationships and breakups | 965 | 18.3 | 26.9 | 47.4 | 7.4 |
| Casual, noncommittal encounters | 867 | 20.6 | 24.6 | 44.6 | 10.1 |
| Sexual content and behaviors | 900 | 16.0 | 22.7 | 52.4 | 8.9 |
| Animated, edited, or promotional content | 758 | 16.5 | 22.0 | 53.8 | 7.7 |

Includes adolescents who rated at least 1 item in the block as seen at least rarely and did not report never seeing that type of content. Sexual content attributed mostly or always to recommended accounts, by profile: Relationship-Socialization, 61.0% (n = 200); Moderate General, 51.0% (n = 298); High Overall, 46.1% (n = 295); Low-Intensity Relationship, 61.0% (n = 100).

### eFigure 1. Bootstrap Community Co-Assignment of Exposure Items

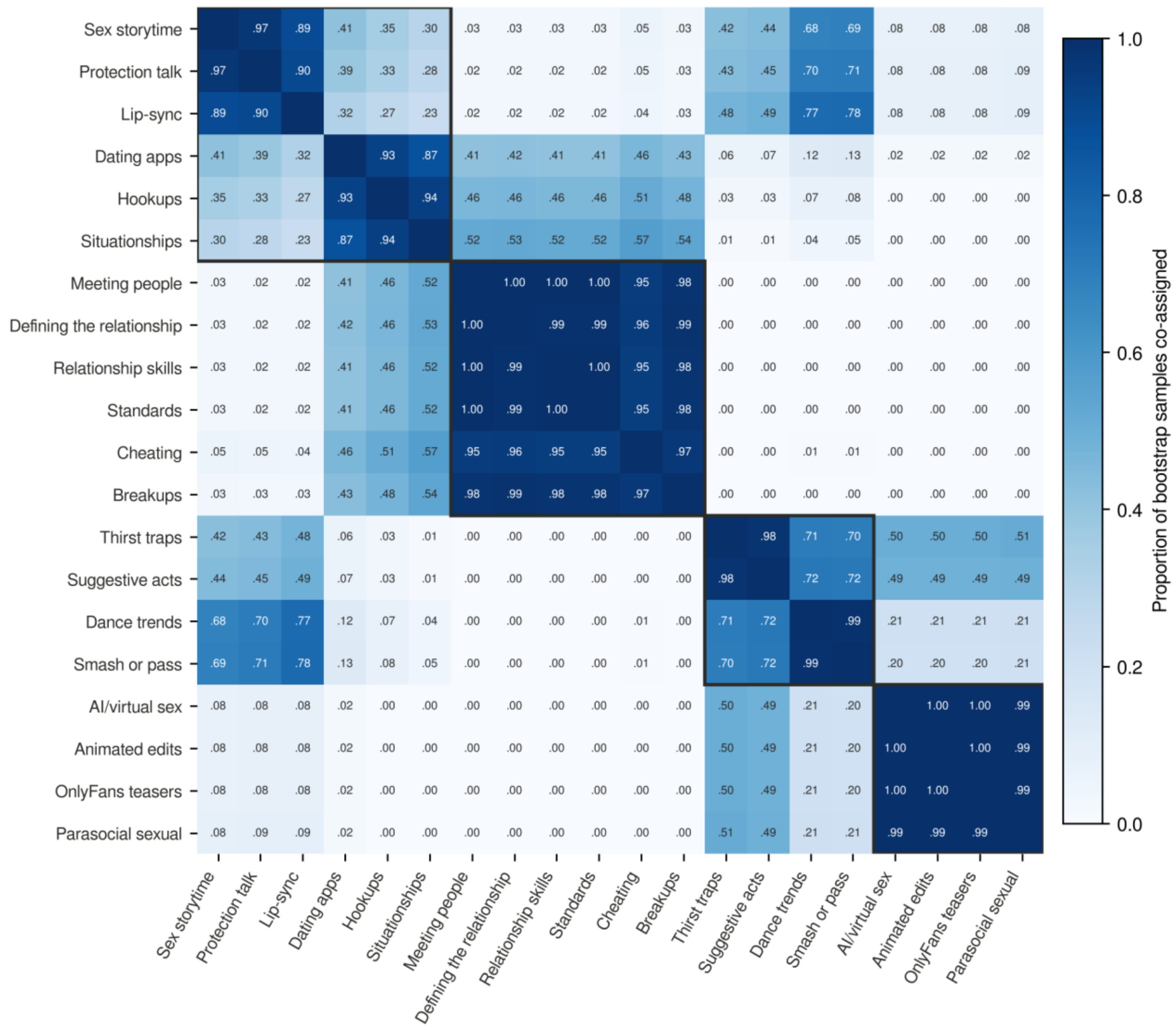


Cells show the proportion of 500 nonparametric bootstrap samples in which each pair of items was assigned to the same Walktrap community. Items are ordered by empirical community (outlined).

#### eFigure 2. Stability of Strength Centrality Under Case-Dropping Bootstrap

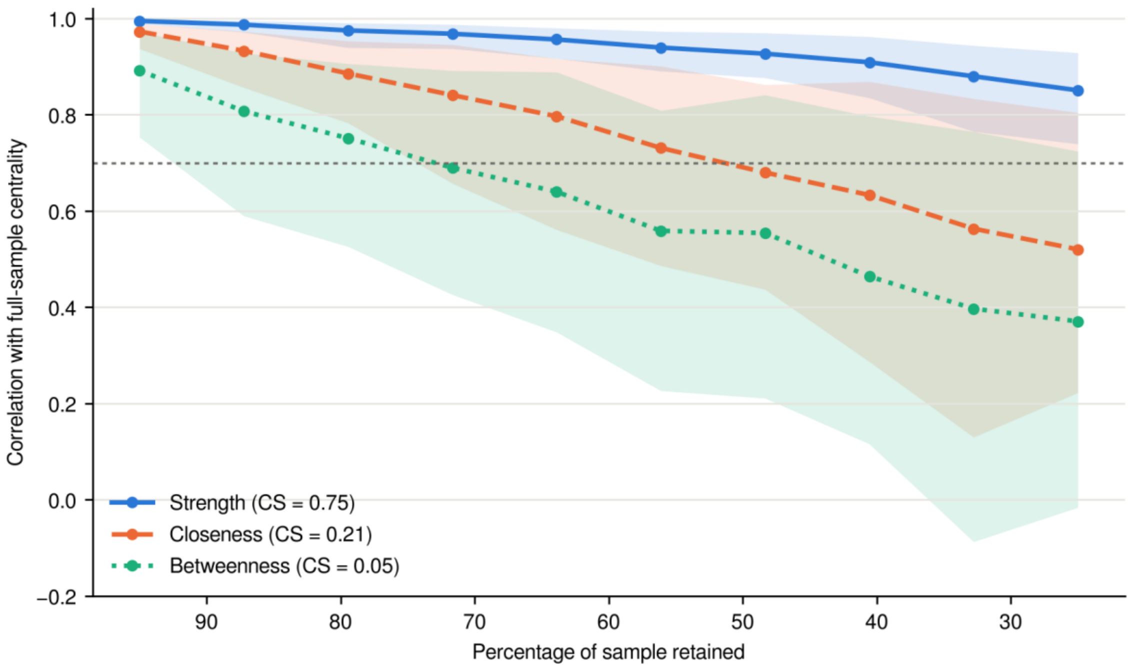


Lines show the mean correlation between centrality indices in subsamples and in the full sample; shaded bands show the 2.5th to 97.5th percentiles across bootstrap samples.
